# HLA-A*26:01 is associated with vertebral endplate bone marrow lesions (Modic changes) in chronic low back pain: a novel immune-genetic link

**DOI:** 10.1101/2025.09.29.25336889

**Authors:** Jan Devan, Tamara Mengis, Pamela Bitterli, Jiamin Zhou, Peter Leary, Po-Hung Wu, Thomas Link, Oliver Distler, Aaron Fields, Stefan Dudli, the REACH Investigators

## Abstract

**Objectives:** Modic changes (MC) are vertebral endplate bone marrow lesions frequently observed in patients with chronic low back pain (CLBP), but their immunogenetic underpinnings remain unknown. We investigated HLA allele associations to uncover immune-mediated mechanisms and potential biomarkers for patient stratification.

**Methods:** We analysed the blood transcriptome from 257 patients with CLBP aged ≥40 years, consisting of 187 patients with MC (types 1–3) and 70 without MC. Bootstrapped LASSO regression identified candidate HLA alleles, which were correlated with MC status using multivariate logistic regression adjusted for demographic covariates. Key findings were validated against US population allele frequencies from 8830 subjects stratified by race and ethnicity.

**Results:** HLA-A*26:01 was strongly associated with MC in CLBP patients (OR = 18.9; FDR = 0.041), and enrichment was independently confirmed by comparison with external population data (OR = 2.15; FDR = 0.008). HLA-DQA1*03:03 was associated with reduced risk of MC (OR = 0.09; FDR = 0.0038). A trend toward positive association was noted for HLA-DRB1*11:01 (OR = 6.0; FDR = 0.069).

**Conclusions:** This is the first study to identify significant associations between HLA and MC. The link to HLA-A*26:01 highlights the significance of CD8^+^ T-cell–mediated immune responses in MC pathobiology. These findings suggest HLA typing may enable personalised treatment strategies in chronic low back pain patients with MC.

**Key messages:**

- What is already known: MC are inflammatory spinal lesions common in CLBP, but their immunogenetic basis is undefined.
- What this study adds: We identify HLA-A*26:01 as a novel risk allele for MC, validated across external population datasets.
- How this may affect research, practice or policy: HLA typing may help stratify patients with MC for targeted therapy or inclusion in immunomodulatory trials.

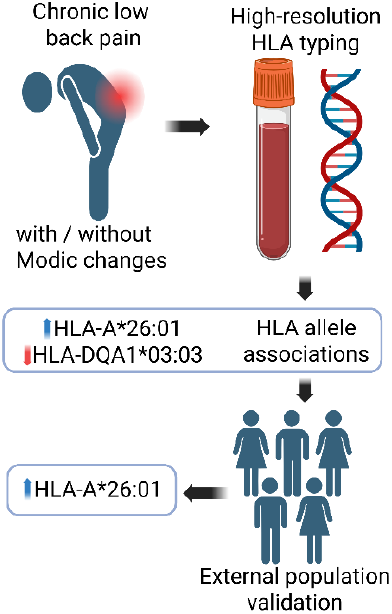

## Introduction

Low back pain is the leading global cause of years lived with disability^1^. While most individuals recover from acute low back pain episodes, approximately one in five adults develops chronic, disabling low back pain^2^. In this population of patients with chronic low back pain (CLBP), Modic changes (MC)—vertebral endplate bone marrow lesions visible on magnetic resonance imaging (MRI)—are frequently observed. On average, MCs are present in 43% of CLBP patients, compared with only 6% in asymptomatic individuals^3–6^ (**Fig. 1**). MCs are categorised into three types based on MRI signal characteristics: type 1 (hypointense T1-weighted [T1w] signal, hyperintense T2w signal), type 2 (hyperintense T1w and T2w signals), and type 3 (hypointense T1w and T2w signals)^3^. MC1 and MC2 are interconvertible, and both may evolve into MC3, suggesting an active and progressive pathological process^7–9^.

**Figure 1.**
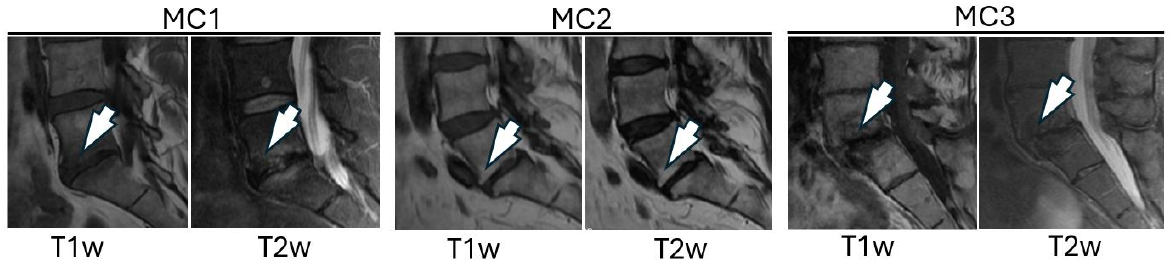
Representative T1- and T2-weighted MRI images illustrating Modic type 1 (MC1), type 2 (MC2), and type 3 (MC3) changes.

Historically, MCs were considered degenerative lesions secondary to mechanical spinal damage. However, recent immunohistological and molecular studies, including our own, have challenged this view. MC lesions exhibit increased vascularisation, immune cell infiltration, and upregulation of inflammatory mediators^8,10,11^. We have shown that MC-associated bone marrow harbours high amounts of activated cytotoxic T cells and antigen-experienced B cells^11–13^. Animal models further confirmed the role of T cells in MC^14,15^. These findings indicate the importance of the adaptive immune system in MC pathogenesis.

HLA molecules are key regulators of antigen presentation and T cell activation. Specific HLA alleles are strongly associated with susceptibility to many immune-mediated diseases^16,17^. We therefore hypothesised that specific HLA alleles may influence susceptibility to MC. In this study, we tested this hypothesis using high-resolution HLA typing in a well-characterised CLBP cohort, and we externally validated key findings using population datasets.

## Methods

### Subjects

For initial examination of HLA allele association with MC, we utilised a subset of participants from The Longitudinal Clinical Cohort for Comprehensive Deep Phenotyping of Chronic Low-Back Pain (cLBP) Adults Study (comeBACK), a multi-site study that is a component of the NIH Back Pain Consortium Research Program (BACPAC)^18,19^. According to the comeBACK study criteria, participants were women and men aged ≥ 18 years, reporting low back pain between the lower posterior margin of the rib cage and the horizontal gluteal fold that had persisted for at least three months and was present on at least half of the days in the preceding six months^18^. In addition, their worst bodily pain had to be located in the lower back, with life expectancy of at least two years, willingness to return for follow-up visits of up to 24 months, and current status as a patient at a participating University of California medical center^18^. All participants had to provide electronic consent^18^. Exclusion criteria included contraindications to MRI (e.g., metallic implants posing safety risk, BMI ≥ 40, claustrophobia), history of spine infection or tumor, inflammatory arthritides (ankylosing spondylitis, rheumatoid arthritis, polymyalgia rheumatica, psoriatic arthritis, lupus), bone-related cancers or metastases, current cancer treatment or treatment within the past 24 months, vertebral fracture in the past six months, cauda equina syndrome or severe leg weakness due to lumbar pathology, current or planned pregnancy within two years, inability to read or write in English, planned enrollment in interfering interventional trials, or any other factor deemed unsuitable by the site principal investigator (e.g., uncontrolled substance or alcohol use)^18^.

Blood samples from 351 CLBP patients were collected. 94 patients were younger than 40 years and were excluded as MC prevalence is rare before the age of 40, and therefore, inclusion of such individuals would risk misclassification of preclinical cases as unaffected^7,20^. Total blood transcriptomic analysis was performed for 257 patients aged 40 years or older, following standardised protocols adapted for BACPAC^21^. Eligibility requires the availability of a clinical MRI^22^.

For validation, we used publicly available data from all published studies reporting HLA-A*26:01 frequency in the U.S. population for White, Black, and Asian racial groups, as listed on the Allele Frequencies Net Database (allelefrequencies.net) and designated as the gold standard. Studies reporting frequencies for racial groups not represented or underrepresented in the initial cohort (defined as <2.5% representation) were excluded.

### Imaging

Lumbar spine MRI was performed on 3-T scanners. Clinical fast spin-echo images with T_1_ and T_2_ weighting were acquired using sagittal and axial prescriptions, as detailed previously^23^. MC were graded on sagittal T1- and fat-saturated T2-weighted images according to established definitions: type 1 MC (MC1) are hypointense on T1- and hyperintense on fat-saturated T2-weighted images, type 2 MC (MC2) are hyperintense on T1- and hypointense on fat-saturated T2-weighted images, whereas type 3 (MC3) are hypointense on both T1- and T2-weighted MRI. Patients showing both MC1 and MC2 at different lumbar levels were classified as MC1, reflecting their greater clinical relevance due to higher metabolic activity, inflammation, and a stronger association with pain^24,25^.

### Blood sample collection

Whole blood samples from patients with varying fasting status were collected in the morning after imaging acquisition in one 2.5 mL PAXgene blood RNA tube for RNA analysis^26^. PAXgene blood RNA tubes were stored upright at room temperature for 2 hours, frozen at -20 °C for 24 hours, and then transferred and stored at -80 °C.

### Transcriptomics and HLA Allele Imputation

RNA was isolated from PAXgene Blood RNA tubes using the RNeasy Mini Kit (Qiagen) and QIAcube Connect (Qiagen). RNA libraries were prepared for each sample with an RNA Integrity Number (RIN)≥ 6 and a concentration of RNA> 5 ng/μL using the TruSeq Stranded mRNA kit (Illumina). The cDNA libraries were sequenced using 100 cycles on a NovaSeq X platform (Illumina) with 100 bp single-end reads to an average sequencing depth of 26 million reads per sample. Sequenced reads were inspected for quality using FastQC and MultiQC. Reads were filtered and trimmed using fastp, and then aligned using STAR to the human reference genome, GRCh38.p13 (GENCODE v42), and was performed on the SUSHI platform^27–31^. HLA alleles were imputed from RNA-seq data using arcasHLA^32^.

### Statistical Analysis

#### HLA Allele Selection via Bootstrapped LASSO Logistic Regression

To identify HLA class I and II alleles associated with the presence of MC (types 1–3) in individuals aged over 40 years, we applied a penalised logistic regression approach using the Least Absolute Shrinkage and Selection Operator (LASSO)^33^ within a bootstrap resampling framework. High-resolution HLA typing data (2-field resolution) were available for 7 loci (HLA-A, -B, -C, -DPB1, -DQA1, -DQB1, -DRB1), from which we extracted standardised 2-field allele notations (e.g., A*26:01). We utilised 2-field HLA allele resolution to balance biological specificity with statistical power. The 2-field designation reflects differences in the amino acid sequence of the HLA protein, which are often functionally relevant to immune recognition and disease association, making them more informative than 1-field alleles that group together multiple functionally distinct variants. Conversely, higher-resolution alleles offer limited additional functional insight in the context of most association studies, while substantially increasing the number of rare alleles, thereby reducing statistical power and interpretability.

Alleles with a frequency below 2.5% in the study population were excluded from the analysis to ensure adequate statistical power. A binary allele matrix was constructed, encoding the presence (1) or absence (0) of each frequent allele per individual, and merged with a binary outcome variable indicating the presence or absence of MC.

To assess the robustness and relative importance of allele predictors, we performed 1000 bootstrap iterations. In each iteration, a LASSO-regularised logistic regression model was fitted to a bootstrap sample of the data using 10-fold cross-validation to select the optimal regularisation parameter (λ). Regression coefficients were extracted at the λ that minimised cross-validated deviance, and the absolute values of coefficients were averaged across all bootstrap iterations to quantify the stability and influence of each allele predictor.

To define a data-driven threshold for variable selection, we ranked all predictors by their average absolute coefficient magnitude. We computed second-order differences to identify an “elbow point” in the ranked coefficient curve corresponding to the point of maximum curvature. Variables with averaged coefficients exceeding this threshold were retained as candidate predictors for downstream interpretation. All results, including ranked coefficients and selected alleles, were exported for documentation and further analysis.

#### Follow-up Logistic Regression and Bootstrap Validation of Selected Alleles

Based on the variable selection results from the bootstrapped LASSO procedure, a follow-up association analysis was conducted focusing on six specific 4-digit HLA alleles (A\**23:01, A\**26:01, DRB1\**12:01, DQA1\**03:03, DRB1\**11:01, DPB1\**13:01) identified as potentially relevant to MC. The dataset was reshaped into a long format (one row per patient–allele pair, allowing a separate logistic regression to be run per allele) and recoded to generate a binary matrix indicating the presence or absence of each selected allele per individual.

For each allele, a multivariable logistic regression model was fitted with MC status as the binary outcome. The presence of the allele served as the main predictor, and models were adjusted for age, sex, BMI, smoking status, ethnicity, and race. To account for potential data separation or sparse data scenarios, Firth’s penalised likelihood logistic regression was applied when any cell in the allele-by-outcome contingency table was zero^34^. Otherwise, standard logistic regression was used.

For each model, regression coefficients, odds ratios (OR), model-based 95% confidence intervals (CI), and Benjamini-Hochberg false discovery rate (FDR)-adjusted p-values were computed^35^. FDR< 0.05 were considered significant. Additionally, a nonparametric bootstrap procedure with 1,000 iterations was used to derive empirical 95% confidence intervals. In each bootstrap iteration, a resampled dataset was drawn with replacement, and the appropriate regression model (standard or Firth) was re-estimated. The 2.5th and 97.5th percentiles of the resulting distribution of log-odds estimates were exponentiated to obtain bootstrap-based CIs. Results include both model-based and bootstrap-based estimates to enhance robustness and interpretability.

#### Validation of HLA-A*26:01 Association Using External Cohorts

To validate the association between the HLA-A*26:01 allele and MC in our cohort, we compared the frequency of the allele against published reference datasets from the United States, curated and listed as gold standard by the Allele Frequency Net Database (AFND). Our cohort dataset was filtered to include only participants with MC (types 1–3) and to retain race and ethnicity groups with a minimum representation of 2.5% to ensure reliable subgroup comparisons. Validation data comprised allele frequency and sample size information extracted from multiple published studies stratified by race and ethnicity. We simulated individual-level genotype data from these summary frequencies via binomial sampling to harmonise with our cohort-level data. The combined dataset included the presence status of alleles, race, ethnicity, and study/source identifiers.

We then employed a generalised linear mixed-effects model with a binomial link function to assess differences in HLA-A*26:01 carrier frequency between our cohort and external validation samples, adjusting for race and ethnicity as fixed effects, and including study/source as a random intercept to account for heterogeneity across studies. Model estimates were expressed as OR with 95% CI. This approach allowed robust assessment of whether the allele frequency in our cohort of patients with MC differed significantly from allele frequencies observed in broader U.S. populations, controlling for demographic variables and study-level variability.

#### Analysis of HLA-A*26:01 frequency worldwide

We utilised HLA-A*26:01 frequency data from all published references listed as gold standard by the AFND. For the countries where more than one dataset was available, the frequency was calculated by pooling all data from all available datasets.

#### Patient and Public Involvement statement

Patients were not involved in the design, conduct, reporting or dissemination of this research.

## Results

### Patient characteristics

Characteristics of CLBP patients with and without MC did not differ, except that there was a higher frequency of never smokers in the CLBP patients without MC (**Tab. 1**). White race was the most common category, encompassing 74.3 % of patients without MC and 77.0 % of patients with MC (p = 0.742). Patients reporting more than one race comprised 1.4 % vs 2.7 % of those without vs. with MC (p = 1.000), and unknown or unreported race was noted in 2.9 % vs. 3.7 % (p = 1.000).

**Table 1:** Characteristics of patients without MC (n = 70) and with MC (n = 187). Values are reported as mean ± standard deviation for continuous variables and as count (percentage) for categorical variables. P-values were derived from Welch’s t-test for continuous measures and Fisher’s exact test for each categorical row.

| Characteristic | CLBP patients without MC | CLBP patients with MC | p-value |
| --- | --- | --- | --- |
| Age (years) | 59.2 ± 11.4 | 61.4 ± 11.0 | 0.174 |
| Body Mass Index (kg/m<sup>2</sup>) | 26.2 ± 4.2 | 26.9 ± 5.1 | 0.251 |
| Sex: Female | 42 (60.0%) | 107 (57.2%) | 0.777 |
| Smoking status: Never smoker | 51 (72.9%) | 109 (58.3%) | 0.043 |
| Smoking status: Former smoker | 17 (24.3%) | 70 (37.4%) | 0.055 |
| Smoking status: Current smoker | 2 (2.9%) | 8 (4.3%) | 0.733 |
| Ethnicity: Hispanic/Latino | 10 (14.3%) | 20 (10.7%) | 0.513 |
| Ethnicity: Not Hispanic/Latino | 59 (84.3%) | 164 (87.7%) | 0.535 |
| Ethnicity: Unknown | 1 (1.4%) | 1 (0.5%) | 0.471 |
| Ethnicity: Prefer not to answer | 0 (0.0%) | 2 (1.1%) | 1 |
| Race: American Indian/Alaska Native | 0 (0.0%) | 1 (0.5%) | 1 |
| Race: Asian | 7 (10.0%) | 20 (10.7%) | 1 |
| Race: Black or African American | 6 (8.6%) | 9 (4.8%) | 0.247 |
| Race: Native Hawaiian or Other Pacific Islander | 2 (2.9%) | 1 (0.5%) | 0.181 |
| Race: White | 52 (74.3%) | 144 (77.0%) | 0.742 |
| Race: More than one race | 1 (1.4%) | 5 (2.7%) | 1 |
| Race: Unknown or not reported | 2 (2.9%) | 7 (3.7%) | 1 |

### Identification of candidate HLA alleles by bootstrapped LASSO regression

Six alleles DRB1\**12:01, DQA1\**05:03, A\**23:01, A\**26:01, DRB1\**11:01 and DPB1\**13:01 were selected by LASSO regression followed by elbow point detection and were retained for downstream modelling (**Fig.2, Tab. S1**).

**Figure 2.**
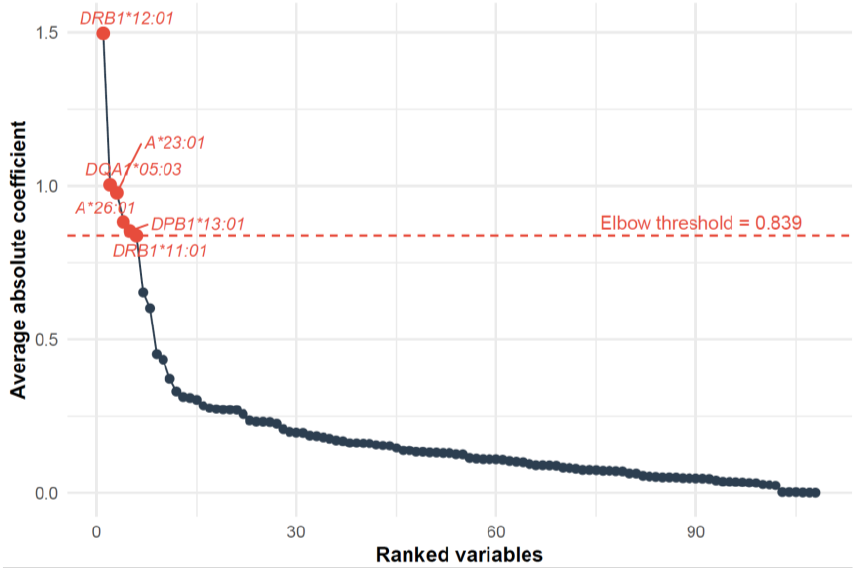
Bootstrap-averaged absolute LASSO coefficients for 3-digit HLA alleles. Points show mean absolute coefficients from 100 bootstraps, ranked from largest to smallest. A dashed red line marks the elbow threshold at 0.839. Six alleles (DRB1*12:01, DQA1*05:03, A*23:01, A*26:01, DRB1*11:01 and DPB1*13:01) lie above this threshold and are highlighted in red.

### Association of Selected HLA Alleles with MC

In follow-up multivariable logistic regression adjusted for demographic and clinical covariates, two of the HLA alleles identified by LASSO selection showed differential associations with MC (**Fig. 3**). Presence of the DQA1*03:03 allele was significantly associated with reduced odds of MC (OR = 0.09, 95% CI: 0.02–0.36; FDR = 0.0038). Conversely, A*26:01 was strongly associated with increased MC risk (OR = 18.9, 95% CI: 1.8–195.2; FDR = 0.041), though the confidence interval was wide. DRB1*11:01 (OR = 6.0, 95% CI: 1.1–32.0; FDR = 0.069) but did not retain statistical significance after multiple testing correction. Alleles DRB1*12:01 (OR = 0.18, 95% CI: 0.02–1.35; FDR = 0.144), A*23:01 and DPB1*13:01 showed weak associations with wide confidence intervals (FDR > 0.1). These findings were further supported by nonparametric bootstrapping (1,000 iterations), with bootstrap-derived confidence intervals similar to model-based estimates (**Tab. S2**).

**Figure 3.**
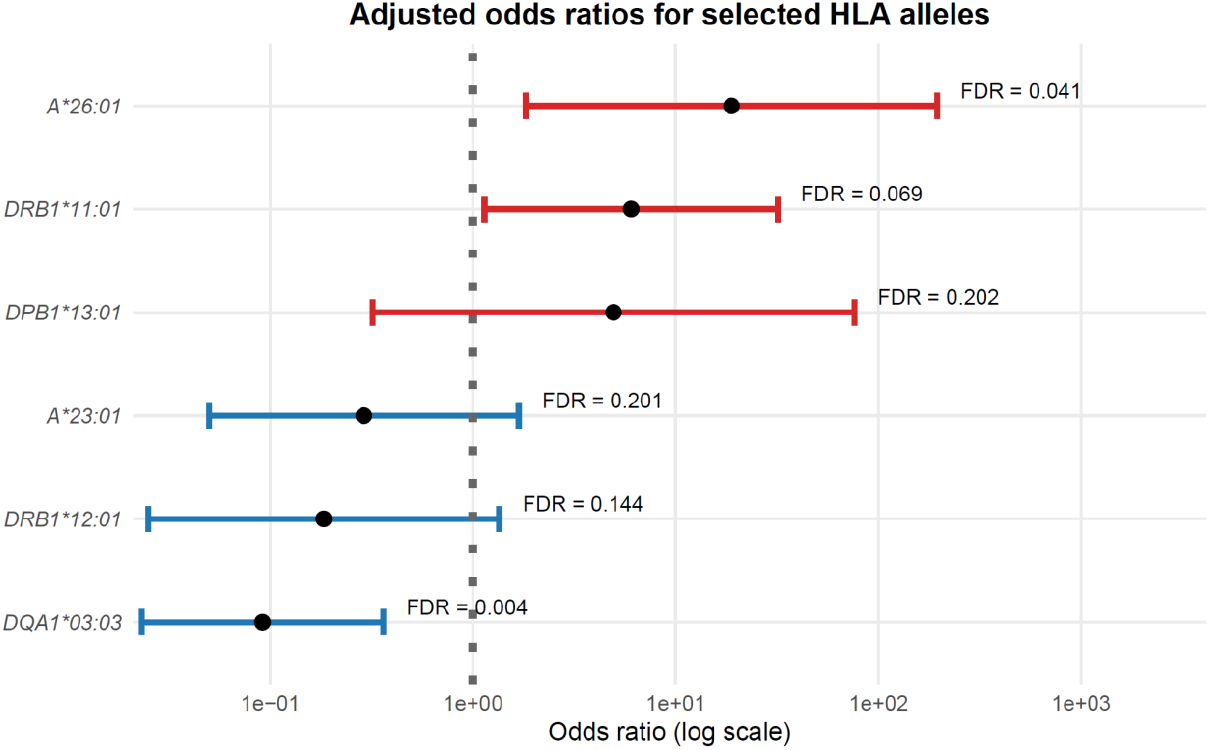
Forest plot of multivariable logistic regression results for selected HLA alleles. Odds ratios (ORs) are shown with 95% model-based confidence intervals. Red intervals indicate alleles with OR > 1, and blue intervals represent OR < 1. A vertical dotted line denotes the null value (OR = 1). False discovery rate–adjusted p-values (FDR) are displayed next to each estimate. Odds ratios are adjusted for age, sex, BMI, smoking status, ethnicity, and race.

### Cross-validation using external population datasets confirms enrichment of HLA-A*26:01 in patients with MC

To assess the broader relevance of the association between HLA-A*26:01 and MC, we compared the allele prevalence in patients with MC in our cohort with allele frequencies reported in studies of subjects from the U.S. population stratified by race and ethnicity (8830 subjects, **Tab. S3**). Mixed-effects logistic regression adjusting for ethnicity, race, and taking studies as random variable confirmed a significantly higher prevalence of HLA-A*26:01 among CLBP patients with MC compared to the general population (OR = 2.15, 95% CI: 1.17–3.42; FDR = 0.008) (**Fig. 4, Tab. S4**). Ethnicity (Hispanic vs. non-Hispanic) had no significant effect on A*26:01 carriage ((OR = 0.90, 95% CI: 0.72–1.20; FDR = 0.537). HLA-A*26:01 was markedly underrepresented among individuals identifying as Black or African American (OR = 0.42, 95% CI: 0.29–0.59; FDR < 0.001), while no statistically significant differences were found for White (OR = 1.49, 95% CI: 0.89–2.49; FDR = 0.143) or Asian individuals (OR = 1.68, 95% CI: 0.91–3.10; FDR = 0.130). A separate race-stratified analysis using binary comparisons (each race vs. all others) similarly confirmed lower odds of HLA-A*26:01 among Black or African American individuals (OR = 0.42, 95% CI: 0.29–0.59; FDR < 0.001), while findings for White and Asian subgroups remained nonsignificant after multiple testing correction. These data indicate a population-specific enrichment of HLA-A*26:01 in CLBP patients with MC relative to the frequency in the general population. Validation for DQA1*03:03 was not possible because of the lack of available data.

**Figure 4.**
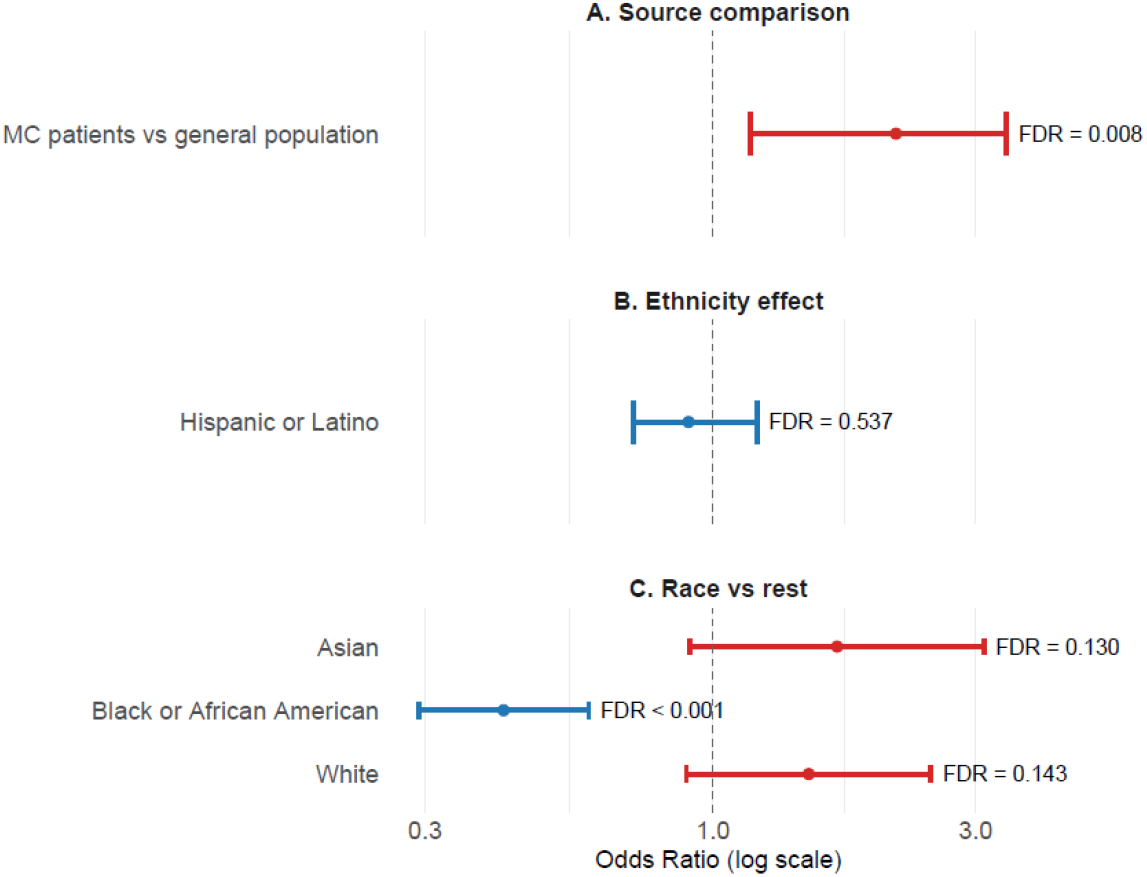
Validation of HLA-A*26:01 association using external population data. Forest plot showing odds ratios (ORs) and 95% confidence intervals from logistic regression models evaluating the prevalence of HLA-A*26:01 across different strata. (A) Comparison of A*26:01 prevalence between Modic change (MC) patients and general population data. (B) Ethnicity-stratified analysis (C) Race-stratified models compared each race category to all others. ORs are plotted on a logarithmic scale; horizontal bars indicate 95% CIs. Red points indicate OR > 1, blue points OR < 1. Dashed vertical line denotes the null value (OR = 1). False discovery rate (FDR)–adjusted p-values are displayed next to each estimate.

### Investigation of the worldwide distribution of HLA-A*26:01

We evaluated 138 datasets from 51 countries (**Tab. S5**). HLA-A*26:01 was most frequent in Oman, Japan and Pakistan, where over 15% of people studied carried the allele (**Tab. S5, Fig. 5**). Between 10-15% of people were bearing the allele in Greece, Bulgaria, Georgia and South Korea. Between 5% and 10% of the population was HLA-A*26:01 positive in studies from Poland, Russia, Iran, Italy, the Cape Verde Islands, Germany, Brazil, Cuba, Madeira (geographically isolated autonomous region of Portugal), Jordan, Sweden, the Azores Islands, China, Guinea-Bissau, Switzerland, and the USA. 2.5%-5% of the population was positive in Portugal, Taiwan, Mexico, Vietnam, Senegal, Singapore, Morocco, Ireland and Cameroon. Less than 2.5% of people carry this allele in Uganda, Papua New Guinea, Burkina Faso, Thailand, Tunisia, Spain, South Africa, India, Kenya, Indonesia, Ecuador, São Tomé Island, Peru, Ghana, and Mali (**Fig. 5**). For all other countries, no data fulfilling our inclusion criteria were available.

**Figure 5.**
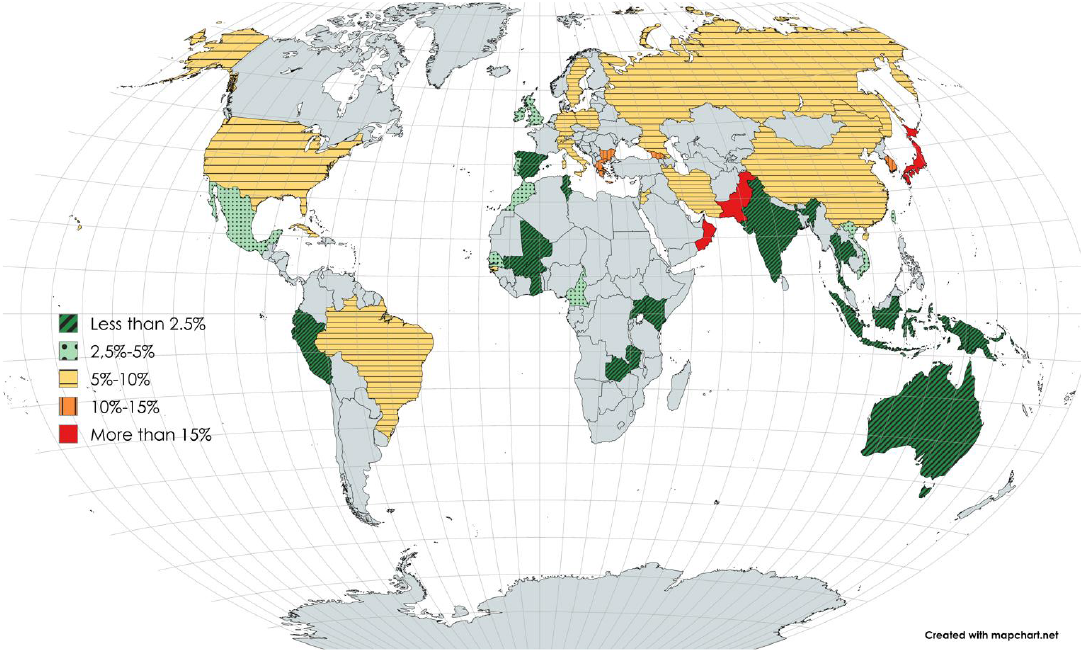
Worldwide distribution of HLA-A*26:01.

## Discussion

We report the first identification of a specific HLA allele associated with MC in patients with CLBP: the enrichment of HLA-A*26:01 among MC-affected CLBP patients. This association was independently confirmed using external population datasets, supporting its robustness. These findings introduce a novel immunogenetic component to the pathophysiology of MCs and suggest that adaptive immune mechanisms—particularly involving antigen presentation to CD8^+^T cells—may contribute to disease susceptibility in a genetically defined subset of patients.

### HLA-A*26:01 is positively associated with MCs in CLBP patients

HLA-A*26:01 has previously been linked to Behçet’s disease and certain lymphoproliferative disorders, but this is the first report connecting it to a musculoskeletal or rheumatic condition^36,37^. The strong association between HLA-A*26:01 and MCs introduces a novel immunogenetic dimension to spinal degenerative pathology. Class I MHC molecules, including HLA-A*26:01, present intracellular peptides to CD8^+^ T-cells, enabling cytotoxic responses against infected cells or cells carrying mutations affecting protein sequences^38^. Unlike HLA-B*27, which is strongly associated with ankylosing spondylitis^39^, HLA-A*26:01 resides on a distinct locus and lacks structural similarity, suggesting independent immune pathways may be involved in MC development.

Endplate damage is hypothesised to expose immune-privileged intervertebral disc components to the bone marrow immune system. In genetically susceptible individuals carrying HLA-A*26:01, this may facilitate aberrant CD8^+^T-cell activation, contributing to inflammation and lesion persistence. These mechanistic insights are supported by our previous identification of cytotoxic T-cell accumulation in MC lesions^11^. Approximately 14% of MC-positive patients in our cohort carry HLA-A*26:01, indicating this is one of several possible pathogenic pathways. The geographic variability of HLA-A*26:01 further underscores the need for replication in diverse cohorts.

### HLA-DQA1*03:03 is negatively associated with MC in CLBP patients

HLA-DQA1*03:03 is associated with the risk of type 1 diabetes^40^ and the risk of late-onset Alzheimer’s disease^41^, but we found it was protective of MC. The lack of data on the frequency of HLA-DQA1*03:03 in U.S. populations prevented similar validation as was done for HLA-A*26:01.

### Clinical and therapeutic implications

Understanding HLA-mediated immune responses in MCs may offer insights into biomarker development and disease stratification. Antibiotic trials for patients with MC1 have shown mixed efficacy^42,43^, likely reflecting the heterogeneous pathobiology of MC. While some MC cases appear to be driven by intradiscal infection or bacterial dysbiosis, other cases might be associated with autoimmune responses^44,45^. Endplate damage associated with MC exposes immune-privileged intervertebral disc tissue to the immune cells of the bone marrow. Alarmins released by tissue damage or from bacterial pathogen-associated molecular patterns from concomitant infection provide an ideal environment for the onset of autoimmunity in susceptible individuals^46,47^. Understanding which MC patients have autoimmune-predominant pathogenesis may be helpful for treatment selection. For example, one might hypothesise that bacteria-associated MC are less likely to benefit from IL-17 inhibitors, as can be seen from the example of other diseases^48–50^. On the other hand, patients with primarily autoimmune MC are hypothesised to benefit from targeted immunomodulatory approaches, akin to TNF or IL-17 blockade in axial spondyloarthritis^51^.

## Conclusion

We identified HLA-A*26:01 as a genetic risk factor for MC—and HLA-DQA1*03:03 as potentially protective against MC—in CLBP patients, which provides strong evidence for an immunogenetic component in MC pathogenesis. These results support the involvement of adaptive immune responses, particularly CD8^+^T-cell activation, in a subset of MC cases. Although it remains to be confirmed, HLA typing may aid in stratifying CLBP patients and tailoring treatment strategies, especially for distinguishing autoimmune-predominant from infection-driven MCs. Further studies are needed to validate these associations in diverse populations and to explore the mechanistic roles of these alleles in MC-related inflammation and tissue damage.

## Limitations

This study has several limitations. First, the sample size is moderate, and no independent, equivalent validation cohort was available. Although we performed cross-validation using external population allele frequency data, replication in larger and fully genotyped cohorts is needed to confirm the association between HLA-A*26:01 and Modic changes (MCs), and to explore the contribution of other HLA alleles.

Second, our analysis used broad racial and ethnic categories, which may obscure important population-specific genetic variation. For instance, Japanese and Thai participants were both classified as “Asian” despite substantial differences in allele frequencies. Moreover, our cohort was predominantly white, highlighting the need for studies in more genetically diverse populations.

Third, the prevalence of smoking—a known risk factor for MC—was higher among individuals with MCs in our cohort. Although smoking status was included as a covariate in all models, residual confounding cannot be entirely excluded.

Finally, this study was conducted in a clinical cohort of patients with CLBP, rather than a population-based sample. This design may influence allele frequency comparisons against general population data, particularly if HLA-A*26:01 is also associated with other CLBP phenotypes. Future studies should assess the frequency and association of HLA-A*26:01 in population-based cohorts, including individuals with asymptomatic MCs, to clarify its role in susceptibility versus symptom expression.

## Data Availability

All data produced in the present study are available upon reasonable request to the authors

## Acknowledgements

We would like to thank the Balgrist Campus for providing generous infrastructure. We also like to acknowledge the Functional Genomic Centre Zurich for their support with sequencing. Research reported in this publication was supported by the National Institute of Arthritis and Musculoskeletal and Skin Diseases of the National Institutes of Health under Award Number U19AR076737. The content is solely the responsibility of the authors and does not necessarily represent the official views of the National Institutes of Health. The Core Center of Patient-centric, Mechanistic Phenotyping in Chronic Low Back (REACH) investigators include the following University of California, San Francisco (unless noted otherwise) personnel in alphabetical order:

Zehra Akkaya, PhD

Prakruthi Amar Kumar, PhD

Jeannie Bailey, PhD

Julia Barylak Sigurd Berven, MD

Andrew Bishara, MD

Dennis M. Black, PhD

Noah Bonnheim, PhD

Atul Butte, MD, PhD

Joel Castellanos, MD (University of California, San Diego)

Jennifer Cummings

Karina Del Rosario, MD

Emilia Demarchis, MD

Sibel Demir-Deviren, MD

Susan K. Ewing, MS

Adam R. Ferguson, PhD

Aaron Fields, PhD

Scott M. Fishman, MD (University of California, Davis)

Sergio Garcia Guerra

Fatemeh Gholi Zadeh Kharrat, PhD

Xiaojie (Summer) Guo

Misung Han, PhD

Trisha Hue, PhD

J. Russell Huie, PhD

C. Anthony Hunt, PhD

Anastasia Keller, PhD

Karim Khattab

Roland Krug, PhD

Gregorji Kurillo, PhD

Feng Lin

Thomas Link, MD, PhD

Jeffrey Lotz, PhD

John Lynch, PhD

Tong Lyu

Rob Matthew, PhD

Wolf Mehling, MD

Esmeralda Mendoza, MPH

Praveen Mummaneni, MD, MBA

Caroline Navy

Conor O’Neill, MD

Jessica Ornowski

Thomas Peterson, PhD

Ananya Rupanagunta (University of California, Berkeley)

Aaron Scheffler, PhD, MS

Shalini Shah, MD (University of California, Irvine)

Irina Strigo, PhD

Naoki Takegami, MD

Abel Torres-Espin, PhD (University of Waterloo)

Salvatore Torrisi, PhD

Sachin Umrao, PhD

Rohit Vashisht, PhD

Joanna Veres

An (Joseph) Vu, PhD

Mark Steven Wallace, MD (University of California, San Diego)

Lucy Ann Wu, MPH

Po-Hung Wu, PhD

Fadel Zeidan, PhD (University of California, San Diego)

Patricia Zheng, MD

Jiamin Zhou, MS

